# Machine learning analysis of population-wide plasma proteins identifies hormonal biomarkers of Parkinson’s Disease

**DOI:** 10.1101/2024.12.21.24313256

**Authors:** Fayzan Chaudhry, Tae Wan Kim, Olivier Elemento, Doron Betel

## Abstract

As the number of Parkinson’s patients is expected to increase with the growth of the aging population there is a growing need to identify new diagnostic markers that can be used cheaply and routinely to monitor the population, stratify patients towards treatment paths and provide new therapeutic leads. Genetic predisposition and familial forms account for only around 10% of PD cases [1] leaving a large fraction of the population with minimal effective markers for identifying high risk individuals. The establishment of population-wide omics and longitudinal health monitoring studies provides an opportunity to apply machine learning approaches on these unbiased cohorts to identify novel PD markers. Here we present the application of three machine learning models to identify protein plasma biomarkers of PD using plasma proteomics measurements from 43,408 UK Biobank subjects as the training and test set and an additional 103 samples from Parkinson’s Progression Markers Initiative (PPMI) as external validation. We identified a group of highly predictive plasma protein markers including known markers such as DDC and CALB2 as well as new markers involved in the JAK-STAT, PI3K-AKT pathways and hormonal signaling. We further demonstrate that these features are well correlated with UPDRS severity scores and stratify these to protective and adversarial features that potentially contribute to the pathogenesis of PD.

## 1. Introduction

Parkinson’s Disease (PD) is the second most common neurodegenerative disease with an estimated incidence of 10 million people worldwide [2]. There is a massive economic burden on individuals, families, and the U.S. government, totaling over 50 billion dollars per year [3] and the number of PD patients is expected to grow with increased longevity of the general population [4]. Parkinson’s disease not only diminishes quality of life but also imposes a substantial societal burden through care giving needs, lost productivity, and high healthcare costs. PD symptoms are marked by tremors, bradykinesia, and other movement-centered symptoms and with disease progression loss of basic movements, and swallowing become difficult. Pathologically, the disease is marked by the death of midbrain dopamine (mDA) neurons in the substantia nigra. This extensive death of mDA neurons in the substantia nigra creates Lewy bodies that impair cell function until death [5] and also releases protein products into serum and cerebrospinal fluid (CSF) [6]. Symptoms often lag disease pathology, and the presymptomatic (prodromal) phase can last around 20 years [7]. The onset of symptoms occur after the majority of dopamine neurons are lost presenting a significant challenge for early intervention and treatment options [8]. A9 neurons are the mDA subtype with the highest loss in PD. Therefore, increase in blood plasma of specific A9 proteins could be strong candidate biomarkers for early detection [9]. Single-cell analyses of A9 neurons lay a foundation that could provide deep insight into PD [10], but are costly and currently cannot be used for diagnostics. Due to the long prodromal phase of PD, early detection is essential for intervention therapy [11, 12]. High throughput proteomic screens allow for fast and cheap identification of impactful proteins. These new minimally invasive screens using blood draws can be applied broadly on aging population to diagnose and provide early targeted treatment whereas CSF or imaging which are costly or too invasive.

Earlier work to understand PD risk centered on genetic information using variations of linear models [13, 14]. Genome-wide association studies (GWAS) are based on the idea that many single nucleotide polymorphisms, which individually have minor associations with PD, would compound and lead to stronger predictive models for developing PD [15]. *Nalls et. al.* has obtained performance from variant-only models trained on the UK Biobank of 0.69 area under the receiver operating curve (AUROC)[16]. Prior work using risk models estimates the probability of developing PD over time, helping to identify individuals at high risk for preventive care. Our current work focuses on developing early diagnostic models which diagnose or categorize individuals as having PD or not, aiding in therapeutic decision-making. More recent approaches like whole genome sequencing, proteomics, or phenotypic evaluation can add significant predictive values. CSF proteomic models haves shown strong classification of PD, but are invasive and carries some risk of complications, which would exclude its use for large-scale screens [17, 18]. More recent methods incorporate machine learning approaches for classification of PD featuring multiomic models spanning transcriptomic to in-depth invasive CSF [19], Sensor [20], and Imaging data[21].

The multiomic models presented here use high throughput proteomic data generated by Olink assays, demographic information and genetic data. The proteomic data is obtained using minimally invasive and low-cost information using just 1 µL of plasma [22]. This information will become more widespread in the future and is ideal for non-invasive combination for screening and understanding the mechanism of disease pathology.

Using proteomic data across large populations can aid the discovery of more disease insights. Neuronal markers that may leak into CSF and then blood have been shown in studies focused on CSF in limited sample sizes [23, 24]. Proteomics data has been collected from Substantia Nigra to identify differences in Pathways between PD case and control [25]. Previous state-of-the-art CSF proteomic model for PD prediction yielded an AUROC for held out sub-cohorts of 0.80, and when combined with CSF and plasma proteomics an AUROC 0.89 [23, 26]. However, CSF and imaging measurements are not routine and typically collected from high risk or diagnosed PD patients which are less applicable for population screens [24]. Furthermore, it is not clear if these biomarkers are correlated with severity of PD, as defined by the unified Parkinson’s disease rating scale (URDPS), such that they can be used to track disease progression.

Past studies of PD biomarkers highlighted a strong inflammatory component in PD [26]. Recent evidence shows inflammation may contribute to the worsening of PD condition and enhance disease progression through reactive oxygen species that mDA neurons are particularly susceptible to [27]. In addition to inflammatory markers, hormonal markers have been shown to have significant correlations with PD progression metrics [28] and lower incidents of PD in women may be partially attributed to protective effect of estrogen [29].

We utilize a deep learning model, a regression model and a SVM model to identify potential novel biomarkers PD. Our results identified nine of our top 20 features from the neural network are hormonally related. We characterized these proteins, such as Prolactin through protein-protein interaction queries as well as through extensive computational analysis and literature reviews. Additionally, we found enrichment for the JAK-STAT pathway which plays a strong role in immunity as well as other functions. We further validated some of our top features identified in the UK Biobank on an independent external validation set. Following this we used top features to determine PD severity through the UPDRS score, canonically accepted as a severity metric for PD. To our knowledge, this is one of the largest population-based proteomic studies for Parkinson’s disease, leveraging cutting-edge machine learning approaches.

Furthermore, we validated the identified biomarkers by correlating them with Parkinson’s disease severity scores, demonstrating their potential utility for tracking disease progression.

## 2. Results

### Study design overview

This study was performed in four stages: data acquisition, data cleaning plus alignment, model architecture tuning, and model interpretation (**Figure 1**). Data were acquired by permission from the UK Biobank and PPMI resources [30, 31]. The two studies used different approaches and technologies for variant detection. The second step consisted of extensive alignment of genomic arrays results, followed by filtering to match the PPMI genomic feature set to that of the UK Biobank. We developed three models, a neural network, support vector machine (SVM), and ridge regression – to classify PD vs control using the proteomics data, genetic variants or combination of both. Training and test of the model was performed on the UK Biobank datasets and external validation was performed on the PPMI data. Model interpretation was performed by extensive literature review of the top features and KEGG pathways to ascertain biological interpretation. To identify the most predictive features, Shapley additive explanations (SHAP) [32] were calculated for the top 85 (12.5% or around 250 top features from top two models) common features between the two top performing models. These features were used in a linear model to predict the PD severity scores defined by Unified Parkinson’s Disease Rating Scale (UPDRS) metric.

**Figure 1:**
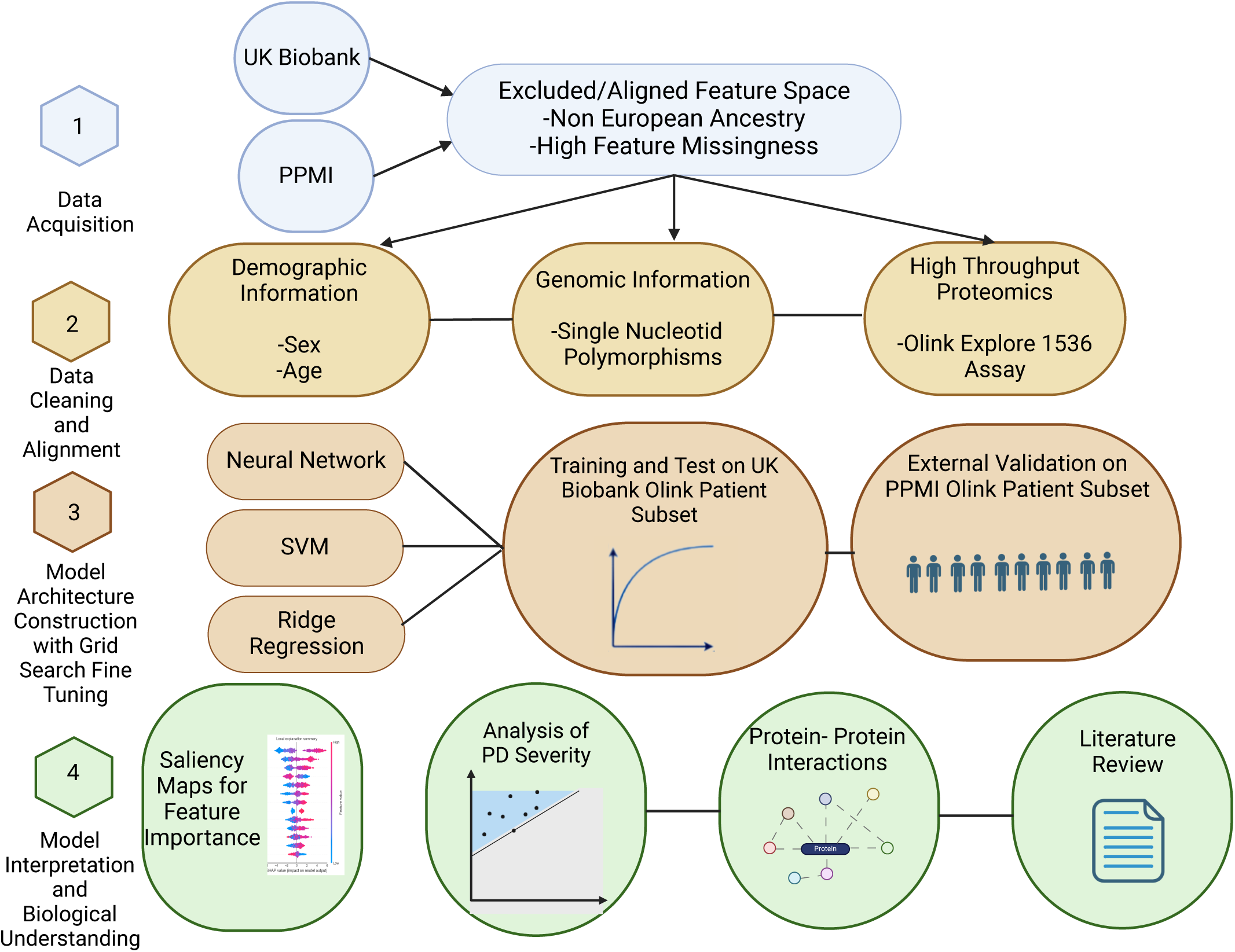
Outline of the study. The study is composed of four steps. In the first step data QC and filtering was performed. In the second step serum proteomic Olink data and SNP polymorphism arrays features from the UK Biobank and PPMI were matched to a common subset. These features were used in a ridge regression, support vector machine and neural network models to predict PD cases in the third step. The last step included biological interpretation of the features and correlation with PD severity scores.

### Dataset populations

In this study we took advantage of two large population cohorts. The first is the UK Biobank that is one of the largest biomedical databases that contains genomics and health related data from half a million UK residents [4, 33]. The second cohort is patient samples from the Parkinson’s Progression Markers Initiative an observational study sponsored by the Michael J. Fox Foundation, aimed at identifying biomarkers to predict, diagnose, and track Parkinson’s disease progression [34]. Both study populations are majority of European ancestry and are skewed towards older patients with a range of 31 to 83 years and median age of 59 years and 60 years for the UK biobank and PPMI, respectively (**Table 1**). Selecting UK Biobank participants that were profiled by the Olink explore 1536 protein panel reduced the number of patients to 43,408. Among those with Olink data 778 patients were diagnosed Parkinson’s disease patients resulting in case-to-control ratio of 2%. The UK Biobank was used predominantly to train and benchmark the predictor algorithms whereas the 103 PPMI cases were used as an independent external validation dataset. The filtered PPMI dataset had a case control ratio higher than the UK biobank of around 24%. This class imbalance is common in a disease setting. Thus, distributions of cases were different which impacted performance on the PPMI validation. This is primarily due to the PPMI data selecting for PD patients while the UK biobank is a population level study. Deep learning models can be ill suited for modeling rare case events due to overfitting [35]. Thus, methods were used to overcome this including dropout, mini-batches, class weighting, and optimized learning rates.

**Table 1:**
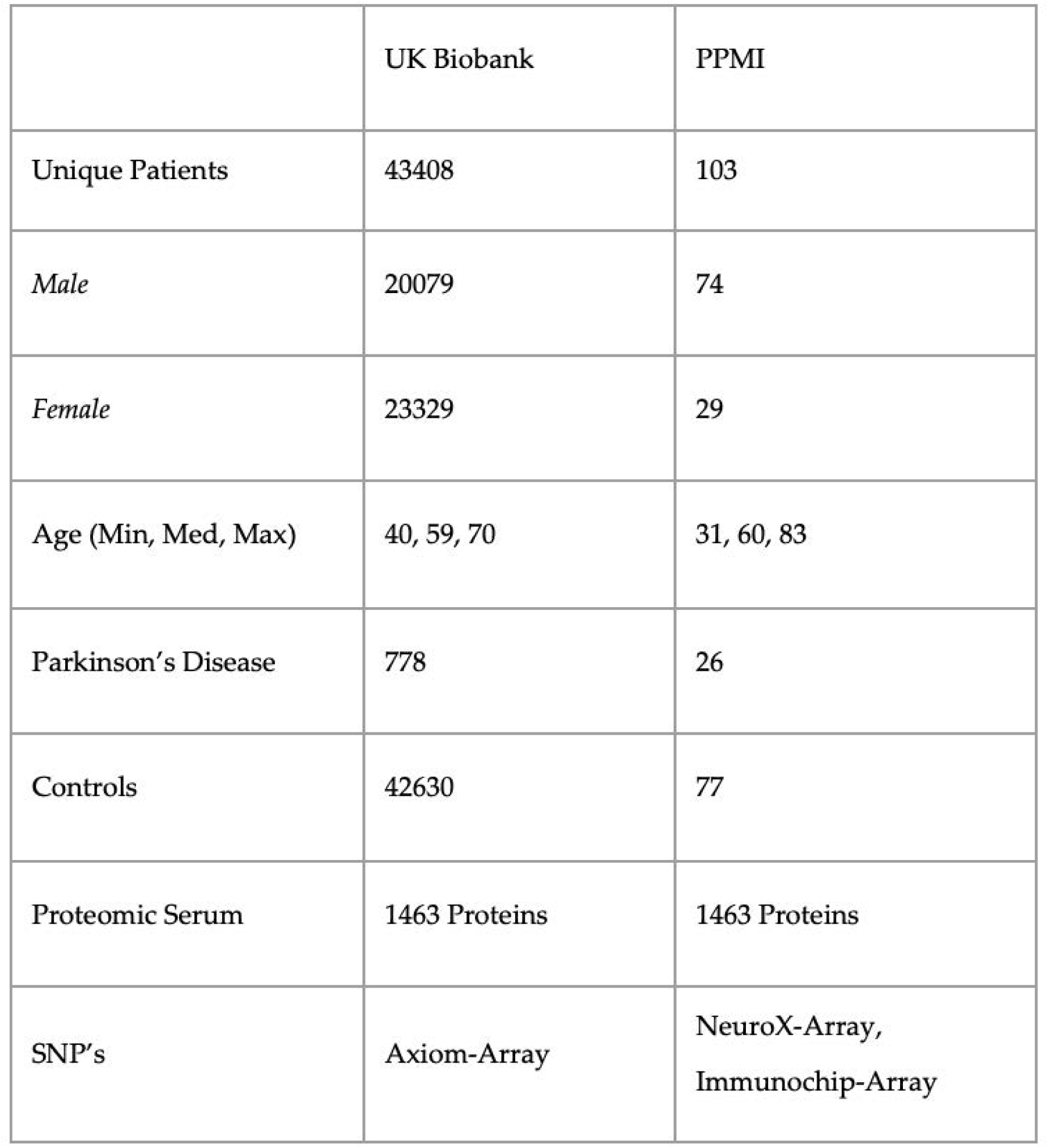
Demographics and feature space of UK Biobank and PPMI cohorts profiled with Olink explore protein panel.

|  | UK Biobank | PPMI |
| --- | --- | --- |
| Unique Patients | 43408 | 103 |
| Male | 20079 | 74 |
| Female | 23329 | 29 |
| Age (Min, Med, Max) | 40, 59, 70 | 31, 60, 83 |
| Parkinson’s Disease | 778 | 26 |
| Controls | 42630 | 77 |
| Proteomic Serum | 1463 Proteins | 1463 Proteins |
| SNP’s | Axiom-Array | NeuroX-Array,<br>ImmunoChip-Array |

### Multimodal proteomic and genetic models

Using the subset of harmonized genomic variants and proteomics data we developed three computational models to classify PD status for individual subjects. We used a neural network, SVM and ridge regression models that input proteomics and variant profiles and produce a probability value for PD diagnosis that is evaluated by ROC analysis of classifier performance (**Figure 1**). Neural Networks (NN) are a general framework for estimating complex, non-linear functions, making them ideal for capturing intricate relationships in high-dimensional data. Support Vector Machines (SVM) identify a dividing hyperplane to maximize class separability, while Ridge Regression is a linear model that uses a penalized linear combination of features to mitigate overfitting. The Area under the cure (AUROC) measures were comparable across all three models on both the training UK biobank and the validation PPMI datasets (**Table 2 & Figure 2**). All three methods had comparable performance of ∼0.77 AUC on the held-out test data from UK Biobank with the NN having highest performance of 0.79 (**Figure 2**). The three models performed similar on the PPMI external validation with AUROC of ∼0.67. The performance drop on the external validation set is potentially due to differences of serum data capture and differences in case-to-control ratios of datasets. Additional factor impacting validation performance could be related to PPMI being drug naïve vs the UK Biobank having a larger number of patients under treatment. When considering sensitivity and specificity, the ridge regression had the highest sensitivity at 0.76 while the SVM had the highest specificity at 0.69. The neural network achieved the best overall balance, with sensitivity and specificity of 0.73 and 0.69. These metrics were both within 3% of the other models, despite a slight reduction in performance on these metrics.

**Figure 2:**
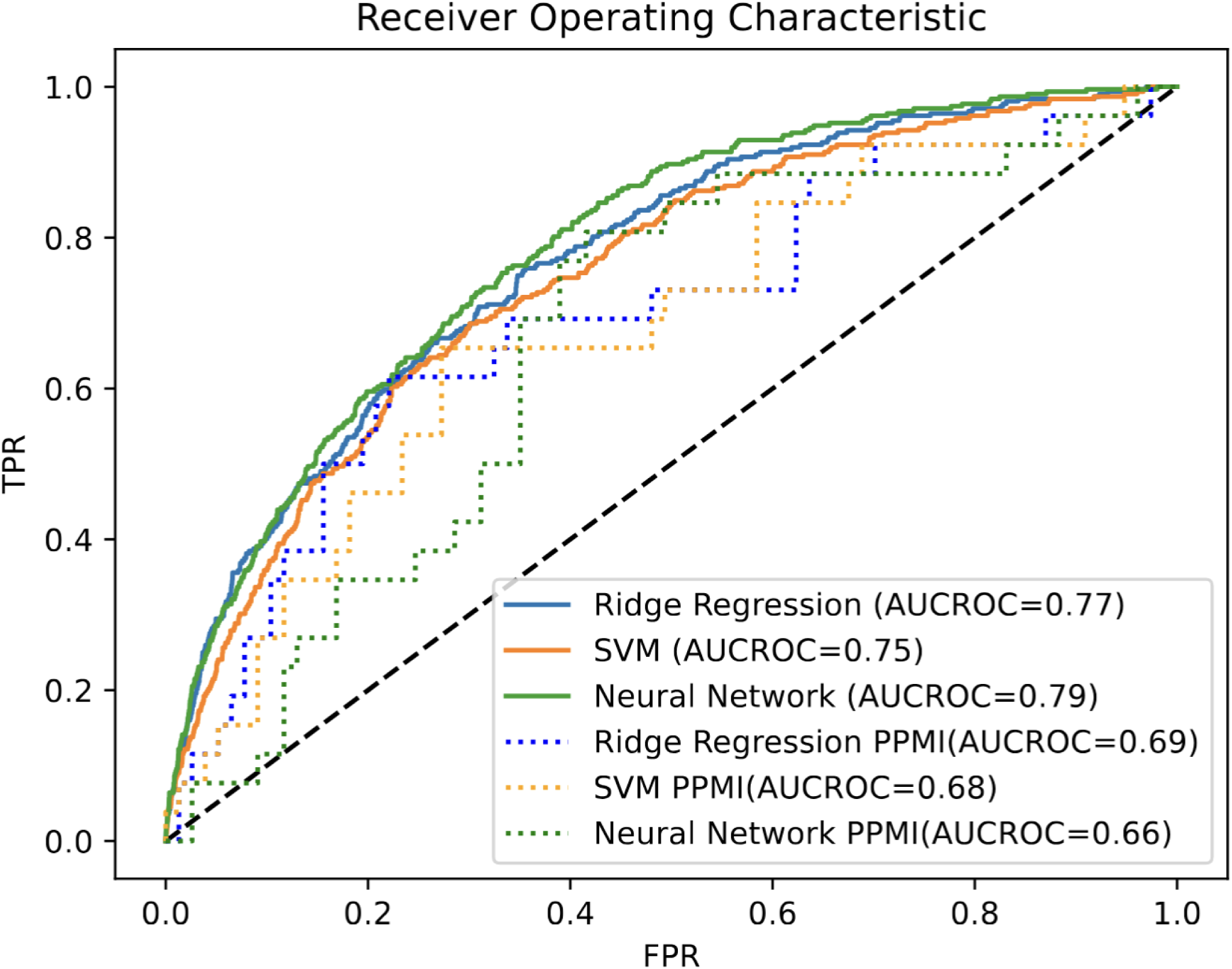
ROC AUC classification performance. AUC of Neural Network, baseline ridge regression and support vector machine on the UK Biobank held-out test set and PPMI validation set.

**Table 2:** Performances of the multiomics prediction models. The three models’ performance was evaluated by AUC, sensitivity, specificity measures for PD classification performance on the UK biobank held-out data as well as AUC performance on the PPMI external validation set.

|  | Ridge Regression | SVM | Neural Network |
| --- | --- | --- | --- |
| AUROC | 0.77 | 0.75 | 0.79 |
| Sensitivity | 0.76 | 0.67 | 0.73 |
| Specificity | 0.64 | 0.70 | 0.69 |
| AUROC PPMI | 0.69 | 0.68 | 0.66 |

### Genetic only model

We created a variant-only model to analyze baseline performance without proteomic data that is akin to a polygenic risk score model. While all three models had reduced performance the neural network AUROC of 0.62 on the training set is significant reduction by over 25% relative to the multimodal model (**Supplementary Table 1**). This was expected as no genetic variants appear in the top 20 feature set in the combined proteomic and SNP neural network (**Figure *3*A**). Although several genetic variants appeared in the top 12.5% of features. External validation performance on the PPMI cases of the variant-only model declined further to random performance of 0.50.

**Figure 3:**
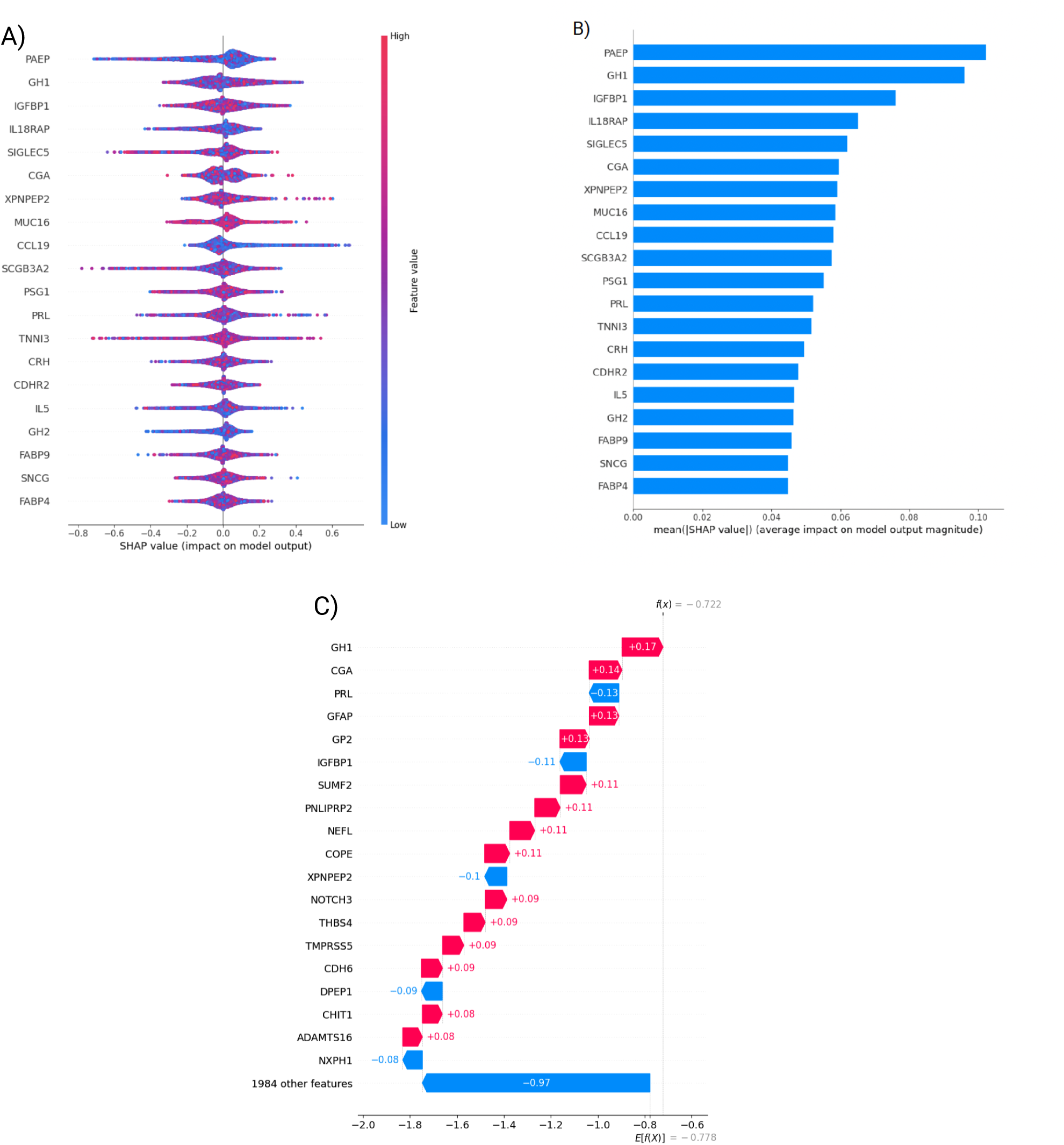
Evaluation of top 20 Neural Network features A) SHAP value distributions of the top 20 neural network features. Each feature is represented by the distribution of SHAP values (i.e. impact on model output) over all test cases. Negative values represent negative weights for PD prediction whereas positive values represent positive contribution to predicting PD. B) Mean absolute SHAP values showing feature importance of the same 20 features. C) A waterfall plot showing SHAP values, contribution to classification result for a single UK biobank participant.

Overall, the use of only genotypic and demographic information mainly sex and age led to substantially reduced prediction performance relative to the multimodal modeling that includes the serum proteomic measurements. This is expected given that these features are typically used for polygenic risk scores to identify individual with high risk, but their genetic penetrance is low. Whereas proteomics data is a more immediate marker for PD state diagnostics.

### Feature evaluation

We used saliency techniques to identify and better understand the most impactful features of both the Olink and SNP for the neural network. We investigated the magnitude of feature impact using the shapley additive explanations (SHAP) score that quantifies features impact on the model performance (**Figure 3A & 3B**). After investigating the top 20 features, we found neuronal specific markers such as DDC and CALB2 that were expected outcomes given that they are related to dopamine function [36] [37]. Unexpectedly, we also identified hormonal markers such as Prolactin and GH1, which are some of the most influential features. SHAP scores allow us to see individual scores from specific patients as well or groups of individuals like all women. For example, in one subject from the UK Biobank study PRL’s negative value indicates a protective contribution whereas GH1’s positive value indicates harmful effect (**Figure 3C**).

Further biological investigation through KEGG analysis of these top markers yielded strong enrichment for Prolactin signaling pathway, JAK-STAT and PI3K-Akt pathways (**Figure 4A**). JAK-STAT and PI3k-Akt pathways were validated in other studies where it was found that PI3k-Akt pathway could be one of the most influential in a rapid progressing PD subtype[38]. The hormonal signaling protein PRL, one of the most widely studied hormones Prolactin, also emerged as a top feature suggesting involvement of hormonal component to PD pathogenesis through activation of JAK-STAT pathways and Prolactin signaling pathways. Additionally, protein-protein interaction enrichment test showed that six of the top features were interrelated hormones that are highly interconnected (p-value < 6.15 e-11, **Figure 4B**) [39]. Our findings suggest that the JAK-STAT and PI3K-AKT pathways may act as central hubs in PD pathology, potentially linking neuroinflammation, hormonal signaling, and neuronal survival.

**Figure 4:**
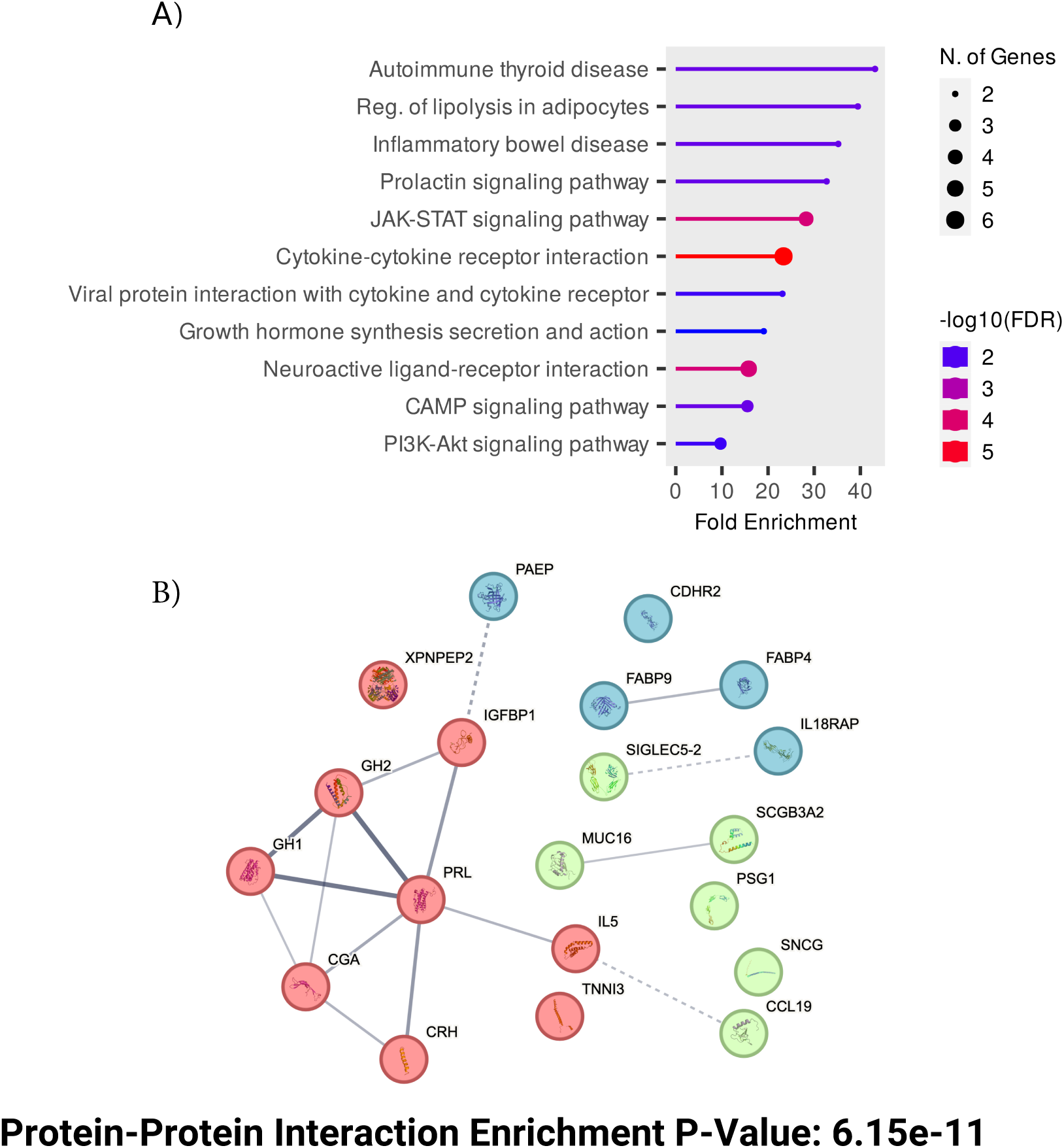
Top features functional analysis (A) KEGG pathway analysis of the top features, based on the genes corresponding to the identified proteins, highlighting key biological pathways involved. (B) Protein-protein interaction (PPI) networks displaying the number of expected interactions of random proteins versus observed interactions. The PPI network exhibits significant enrichment with a p-value of 6.15e-11. Three distinct clusters are shown in different colors. With the thickness of connections denoting strength of connectivity across experimental data and literature data.

### Feature correlation with PD severity score

We next investigated the shared features that the different models identified as significant to identify the core biological pathways that are predicative of PD. We subset the top 85 (12.5%) features that were common across the ridge regression and neural net models, excluding the SVM model due to slightly lower performance. These 85 common features were used to construct a linear regression to predict UPDRS [40], which is the accepted severity score for PD among physicians (**Figure 5**). We found that these top features are well correlated with UPRDS values in the PPMI dataset (R^2^ = 0.86). Several SNP’s were included in this top feature set as highly predictive of PD that were previously validated in the literature. These including rs12951632, rs11158026, and rs10748818, indicating their established association with Parkinson’s disease and further supporting their potential role in disease susceptibility and progression [41, 42]. Two of the variants, rs12951632 and rs11158026, were identified in a related genome wide association studies as variants that are strongly associated with PD. The third variant, rs10748818, was previously identified as causing changes in the GBF1 gene that is a critical factor in PD pathogenesis [43].

**Figure 5:**
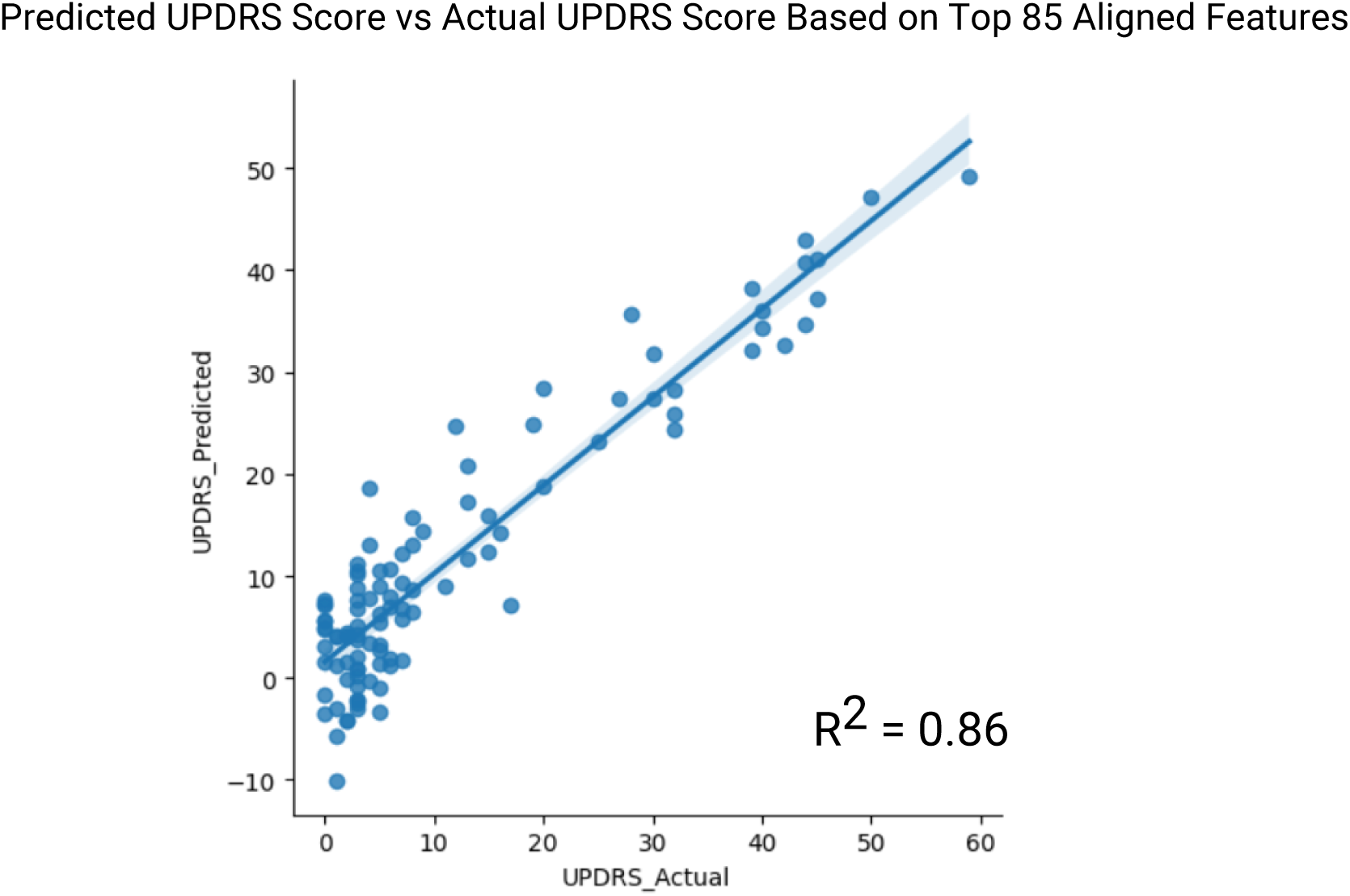
Linear model of URPDS scores. Top 85 aligned features of the neural network and the ridge regression were used in linear regression to predict UPDRS score on PPMI validation set (R2 = 0.86).

#### Feature signatures associated with increased PD

Next, we were interested in investigating the contributions of the features to likelihood of PD. K-Nearest Neighbors clustering of the PPMI cohort using the shared 85 features identified one group of participants that had high PD vs control ratio (cluster 1, **Figure 6A**). This cluster is strongly distinct form other clusters by a protein signature that is driven by 14 proteins that are negatively correlated with the PD (**Figure 6B**). One of these proteins is FABP5, a fatty acid binding protein, which was recentrly identified as a PD marker in an independent proteomics study of 99 PD patients [44]. In addition, a similar study by *Hallqvist et. al.* used mass spectrometry approach to identify protein markers from CSF and serum that predict PD in the prodromal phase among a small cohort [44]. While not all significant proteins from their study overlapped with the protein measured in the Olink assay, five overlapping proteins (CST3, DKK3, FABP5, PTGDS, and VCAM1) were found as significant PD markers in our study (p-value < 0.10, by t-test).

**Figure 6:**
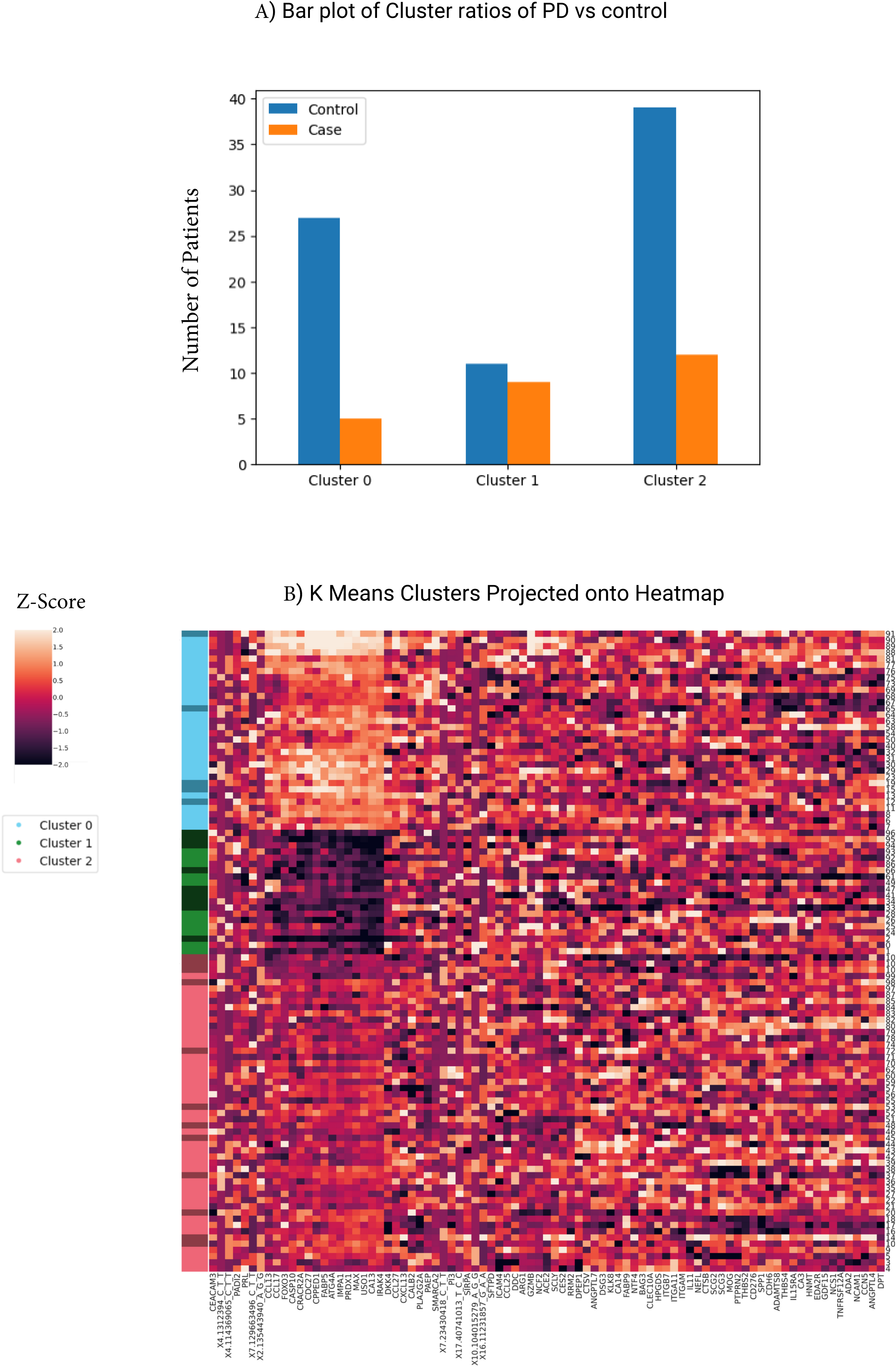
Feature clusters identify protective PD proteins (A) KNN clustering of top 85 features identified three predominant clusters. Bar plot highlighting the enrichment of PD relative to controls in Cluster 1. (B) Heatmap of protein levels (Z-scores) expression in the PPMI datasets. Left side color strip indicates cluster assignment (cluster 0 is blue, cluster 1 is green and cluster 2 is red) where darker green, blue or red indicate PD cases while lighter colors indicate controls. Proteins are clustered to identify cluster signatures.

### Feature correlations

To gain insights into the cross-feature correlation structure between the predictive features we performed pairwise correlation analysis with UPDRS scores (**Supplementary figure 1**). Two distinct highly correlated protein groups were identified. The first group of proteins characterized show cytokine-cytokine enrichment as well as involvement in PI3K-AKT signaling pathway (**Supplementary figure 2A**). This signature has a strong immune and inflammatory component which has been a hallmark of PD and every feature was positivley correlated with UPDRS scores. The second group (marked in blue, **Supplementary figure 2A**) is associated with MAPK signaling pathway as well as other apoptotic pathways (**Supplementary figure 2B**). MAPK signaling has been strongly implicated in PD with connections to high penetrance variants such as LRRK2 [45]. This signature is similar to the downregulated signature assoicated with increase PD cases and is inversly correlated with UPDRS scores suggesting a protective role in PD pathogenesis (**Figure 6B**).

The most correlated protein with UPDRS severity is ANGPTL4 (Pearson = 0.57), a lipid homeostasis modulator where elevated levels were recently associated with neurodegeneration [46]. Conversely, the least correlated protein (Pearson = −0.56) is HPGDS, an enzyme that is involved in proteinoids synthesis and anti-inflammatory response [47]. Overall, feature clustering on the PPMI dataset the show strong protein signatures associated with PD cases and UPDRS scores. The high overlap between the cluster signature and the signature in the feature-feature correlation is possibly a core protein signature that can inform future studies to better understand PD mechanisms.

## 3. Discussion

Our PD prediction algorithms from plasma proteomics confirmed several established biomarkers as well as potentially novel biomarkers that warrant further investigation. The top features in both the ridge regression and neural network models are DDC and CALB2 which have previously been implicated in PD [48, 49]. DDC is involved in dopamine synthesis [50] and is directly related to massive cell death of dopamine neurons in the striatum [51]. CALB2 is heavily featured in single-cell analysis of PD as a key marker for A10 dopamine neurons as compared to A9 dopamine neurons [49]. Ridge regression coefficients show CALB2 as negatively associated with PD which is consistent with its expression in A10 mDA cells that are mostly spared from cell death during PD progression [52].

In our study, several other less direct markers, like hormonal components, were significantly correlated with PD incidents. These hormonal features, especially Prolactin, were significantly correlated in multiple models and cluster analysis. Our findings illustrate a strong hormonal component, with six of the top 20 proteins identified by the neural network being hormonally related through heavy neurodevelopmental involvement and links to apoptotic functions. Further evidence from protein interaction analysis identified these six proteins as closely functionally related (p-value < 6.15e-11, **Figure 4B**).

Another significant marker, Human Growth Hormone (GH1) has several known functions ranging from growth of various tissues to neurogenesis [53]. Notably, GH1 has been used to treat amyotrophic lateral sclerosis in vitro as well as in mouse models demonstrating strong neurological protection [54]. This is consistent with our findings where GH1 is enriched in the control group relative to PD cases making it a potentially protective marker. Prolactin produced by lactotrophs in the pituitary gland is another hormone that could be critical to understanding PD [55]. Prolactin (PRL) is known for its multifaceted impacts including neuroinflammation, which is part of the pathogenesis of PD [56] and reducing neuroinflammation through modulation of PRL may offer a new therapeutic path for PD. Studies have also demonstrated that prolactin release is regulated by dopamine [57] and therefore, loss of PRL may indicate loss of dopamine.

One common inflammatory model used to study neuroinflammation in mouse models is through intranasal lipopolysaccharide injections that cause widespread inflammation throughout the brain and allow researchers to investigate these the effect of inflammation on neurodegeneration. These inflammatory models lead to selective dopamine SN loss, striatal dopamine depletion, and increased alpha synuclein in the SN [58]. This loss suggests that inflammation in the brain preferentially targets dopamine rich regions than other areas of the brain and points to possible association of PD pathogenesis with inflammation. Furthermore, it was shown that in this lipopolysaccharide model PRL is part of a full-autocrine loop that enhances inflammatory response that may contribute to further worsening PD through enhanced inflammation [59].

PRL and GH1 play a strong role in the brain with widespread expression. This role especially extends past simply neurodevelopmental. PRL and GH1 are highly expressed in the cerebellum. (**Supplementary Figure 3A)**. Findings of cerebellar circuitry alteration in PD highlight the importance the cerebellum play in PD progression and disease pathology[60]. The cerebellum is perhaps more involved in PD pathology than previously thought and may serve as an understudied area that contains biomarkers indicative of PD.

We performed KEGG Pathway enrichment analysis with the top 20 neural network features which further supports the potential role of JAK STAT and AKT pathways in PD pathogenesis. Jak2/Stat5 are commonly linked to developmental embryogenesis [61]. These pathways are likely linked to PRL and GH1 activities as both receptor bindings can lead JAK2 activation [61] (**Supplementary Figure 3B**).

While our study focused on PD, it represents a new framework to investigate other diseases through multiomic integration of proteomics and variants in the UK Biobanks and other public data resources [30, 62, 63].

We have laid the groundwork to continue building on this multiomic model to provide an analysis framework to integrate proteomic Olink data and variant data in the UK biobank. This approach can identify new association with other neurodegenerative diseases. For example, through phenotype-gene enrichment analysis we found several phenotypic PD features associated with height which has some suggestive correlation with PD (**Supplementary figure 4**) [64, 65]. With the increase of Olink data in the UK biobank such new insights into the mechanisms of these diseases can now be investigated.

Plasma proteomics emerges as a powerful, minimally invasive alternative to cerebrospinal fluid-based biomarkers, achieving comparable classification performance while offering broader accessibility for large-scale screening. By leveraging machine learning models, we identified biomarkers including hormonal markers such as prolactin and GH1 that suggest an underexplored hormonal axis in PD pathogenesis. Additional, pathway analysis implicating the JAK-STAT and PI3K-AKT pathways in linking neuroinflammation, hormonal signaling, and neuronal survival. These findings not only demonstrate the feasibility of integrating plasma proteomics into cost-effective, population-level screening programs for early diagnosis and intervention but also provide a framework that can be extended to investigate biomarkers for other neurodegenerative diseases.

## 4. Materials and Methods

### Study Population

The study population consisted of two groups from the UK Biobank cohort and Parkinson’s progressive marker initiative cohort [30, 34]. The UK Biobank is a large-scale biomedical database containing de-identified information of over 500,000 patients including genotyping as well as serum analysis. Diagnosis of Parkinson’s disease was made by International Classification of Diseases coded as ICD-9 and ICD-10 codes. Single nucleotide polymorphisms were taken from the Axiom Array. The UK Biobank after processing for missingness and filtering for Olink data consisted of 48,408 patients: 23,329 female and 20,079 males with median age of 59. The PPMI dataset is an international consortium across 12 countries that is primarily sponsored by Michael J. Fox Foundation [34]. PPMI uses 2 arrays for SNP’s including Immunochip and NeuroX specially designed for PD After processing there are around 103 individuals consisting of 29 female and 74 males with a median age of 60. Diagnosis of PD was made with the addition of DAT imaging to enhance accuracy [34].

We subset these datasets for participants that had Olink serum data collected. This substantially limited the number of participants. Both datasets used Olink Explore 1536 so feature alignment was possible. 1484 proteins were measured from serum via proximity extension assay (PEA) high-multiplex immunoassay. Olink Explore 1536 contains 4 separate panels Oncology, Cardiometabolic, Inflammation, and Neurology [66]. The assay works by double antibody binding to proteins leading to DNA hybridization which can be PCR amplified, sequenced, and quantified. Proprietary normalization of these values provides protein quantification allowing comparison across individuals and studies.

### Variant Preprocessing and alignment

Genomic variants were preprocessed using PLINK 2.0 software, a genome association toolkit [67]. We aligned the variant arrays in both arrays by position, major allele, and minor allele to Genome Reference Consortium Human Build 37. The UK Biobank used the Axiom array, while the PPMI dataset used the Immunochip and NeuroX array. We filtered for the top 324 overlapping variants and used additional 311 variants identified in the *Kim .J.J. et al*. study[68]. This proprietary dataset allowed for more power leading to identification of associated variants. In total, 635 filtered variants with overlap across both the UK biobank and PPMI validation set were used in our prediction models.

### Study Design

The UK biobank was used for training and test, and PPMI data was used an independent external test to measure classifier performance. Patients were filtered for European ancestry and missing features. In addition, features were filtered for missing values. The resulting multiomic dataset combined demographic data, genomic data and proteomic data. We used several baseline models such as a ridge regression which performs L2 penalization to minimize coefficients [69]. Additionally, a SVM with a linear kernel was used as a second baseline model. For training and test, a split of 60% and 40% was used, respectively. The ratio of PD cases to control was maintained in training and test splits. The genetic single nucleotide polymorphism only model utilized the same split of 60% and 40% for the training and test dataset. Ridge regression and SVM were tuned by grid search to identify the best model. Both utilized classes weighting to address imbalance common with medical data.

The neural network architecture is a hidden 2-layer neural network. In this fully connected neural network the size of the first hidden layer is 120 and second hidden layer is 40. Activation was done by leaky ReLU. A sigmoid function was then applied for the final output layer binary prediction value. The model uses binary cross entropy (BCE) as the loss function that assigns a larger error for mislabeled cases as compared to mean square loss, which is more appropriate for training data with relatively low fraction of positive cases.

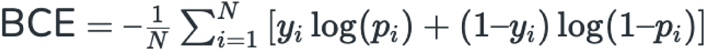

Where N is the number of samples, y_i_ is the label (i.e. PD or control) and p_i_ is the predicted probability of y_i_. Several steps were taken to deal with imbalanced cases, including class weights and model architecture adjustments. Dropout was utilized to address overfitting which was emboldened by imbalanced cases. Furthermore, a large batch size was used to give more positive PD cases for each stochastic gradient descent step using Adam optimizer.

### Predicting UPRDS scores

A linear model was built using the 85 neural network and ridge regression common features on the PPMI dataset for UPDRS score. These 85 features were used in a regression to predict UPDRS scores to further validate our top features. Sample clustering was performed through both ward hierarchical clusters and K-means where K=3 was selected silhouette score and cluster inspection.

### Feature Correlation Analysis

Feature-feature correlation analysis was performed on these top 85 features using Pearson correlation. The UPDRS score was included in these correlation calculations and hierarchal clustering of the correlation values (Supplement Figure 1).

### Model Interpretation

Feature weights on the ridge regression was done through the coefficients placed on the optimized model. The neural network used a game theoretic approach to feature importance called SHAP scores [32]. These scores create a powerset of features to determine marginal contribution of each feature. Mean absolute SHAP scores were taken across all combinations of models on the testing set to determine the feature importance. The distribution was visualized through a bee swarm plot. Additionally,

Phenotype-gene enrichment analysis was done using to detect statistically significant phenotypes from our top 20 features.

## Data Availability

UK Biobank and PPMI genomic and phenotypic data is restricted and requires prior approval for access and therefore cannot be provided in this publication.

## Code availability

All code written and used for the above analysis and figures is freely available at https://github.com/chaudhry123/PD_Biomarkers_Project.

## Acknowledgments

We thank members of the Betel lab for helpful discussion as well as members of Lorenz Studer and O.E groups for helpful discussion and data access. D.B. and F.C. were supported by R01 NS118067 from NIA. F.C is supported by NIH grant T32 GM132038. T.W.K. was supported from a National Research Foundation of Korea (NRF) grant funded by the Korea Government (MSIT) (no. RS-2024-00351442). Data used in the preparation of this article was obtained on 2023-01-31 from the Tier 1 Parkinson’s Progression Markers Initiative (PPMI) database (www.ppmi-info.org/access-data-specimens/download-data), RRID:SCR_006431. For up-to-date information on the study, visit www.ppmi-info.org. PPMI – a public-private partnership – is funded by the Michael J. Fox Foundation for Parkinson’s Research, and funding partners; including Aligning Science Across Parkinson’s (ASAP) initiative. Other partners include a consortium of industry players, non-profit organizations and private individuals. This research has been conducted using the UK Biobank Resource under Application Number 47137

## Author Contributions

FC and DB performed data analysis, data processing, and model development. OE participated in discussions to conceptualize the idea, provided access to data, and contributed to the development of the manuscript. TWK provided biological interpretation of the results and assisted in manuscript preparation.

## Supplementary Material

**Supplement Table 1:** Performances of PD prediction models using genetic variants and demographic data. Like table 1, the performance of the three models using SNPs variants was evaluated by AUC, sensitivity, specificity measures for PD classification performance on the UK biobank held-out data as well as AUC performance on the PPMI external validation set.

|  | Ridge<br>Regression | SVM | Neural<br>Network |
| --- | --- | --- | --- |
| <u>AUROC</u><br>Test | 0.70 | 0.70 | 0.62 |
| Sensitivity | 0.72 | 0.59 | 0.37 |
| Specificity | 0.55 | 0.68 | 0.78 |
| AUROC<br>PPMI | 0.55 | 0.45 | 0.51 |

**Supplement Figure 1:**
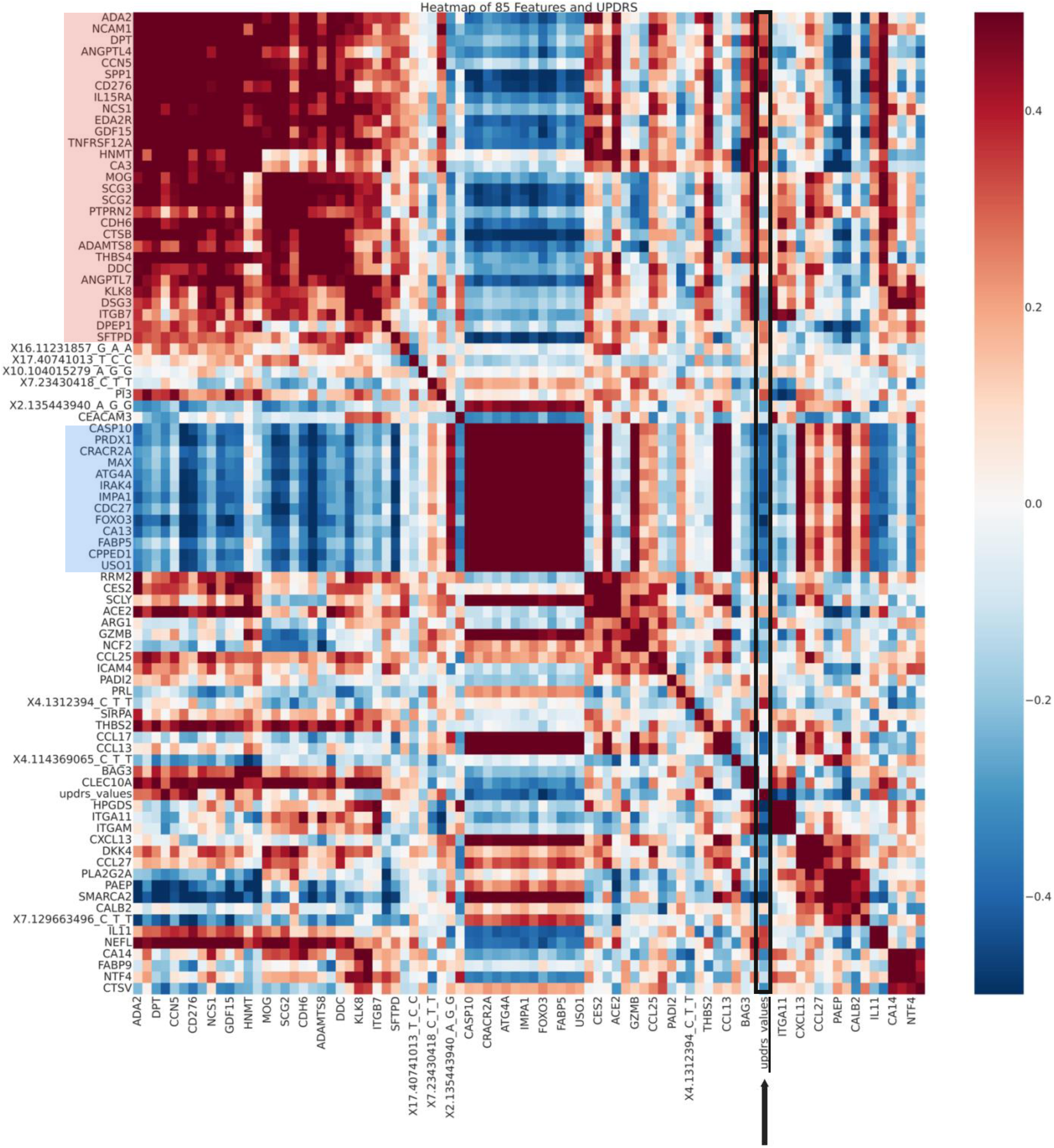
Feature correlation. The Pearson correlation heatmap between 85 selected features and the UPDRS score, with color intensity indicating the strength and direction of the correlations. Strong correlations are marked by darker shades, while lighter shades represent weaker correlations, highlighting potential patterns and associations relevant to Parkinson’s severity. Two main feature groups are strongly correlated (in red) and anticorrelated (in blue) with UPDRS scores (marked by black arrow). Note that due to space constrains every second feature is labeled on the vertical axis.

**Supplementary Figure 2:**
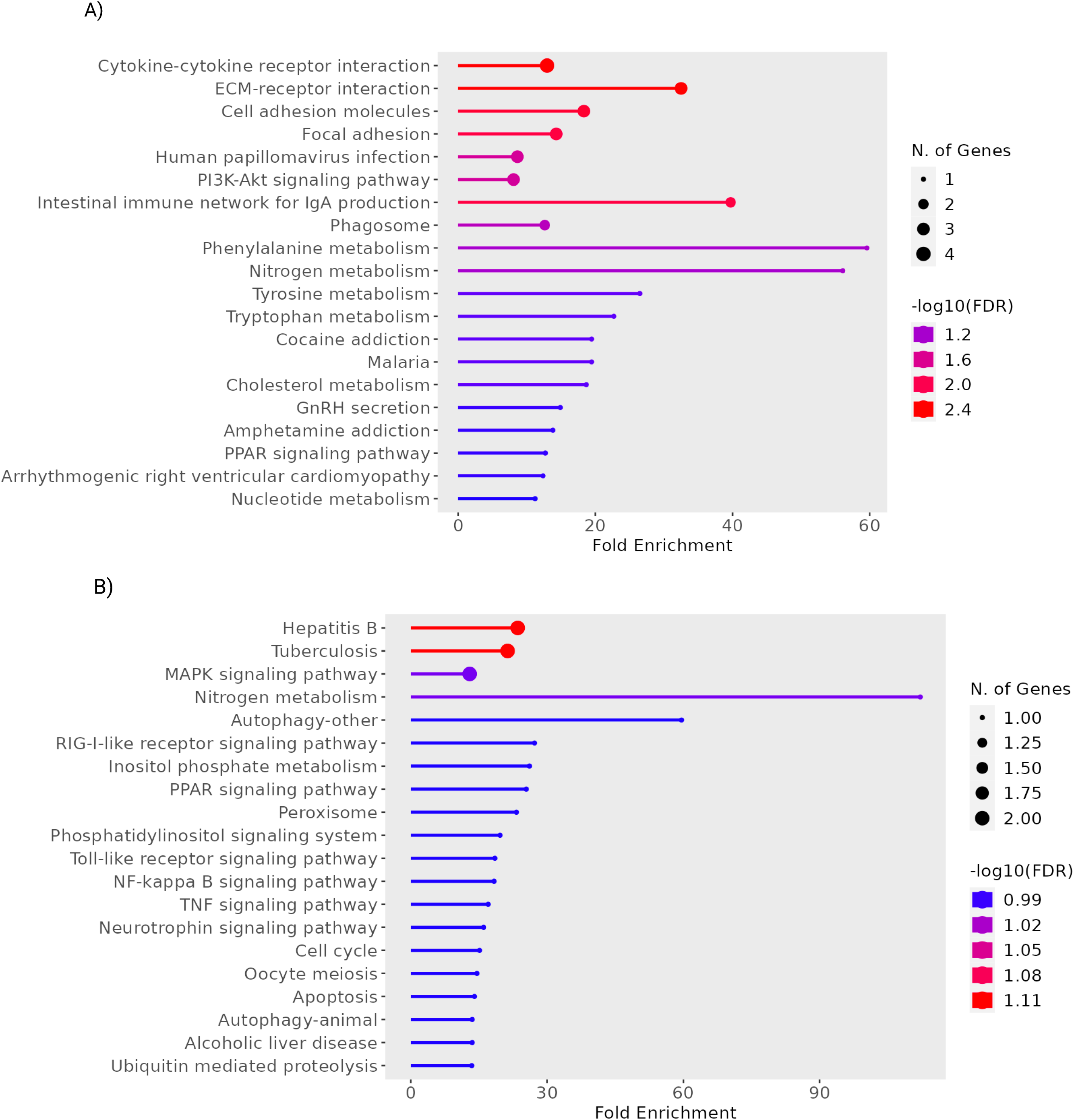
Pathway enrichment of UPDRS correlated features. A) KEGG analysis of top UPDRS correlated features (red in group in Supplementary Figure 1) highlight enrichment in the cytokine-cytokine receptor interaction and PI3K-AKT signaling pathways. B) Similarly, enrichment analysis of top UPDRS anticorrelated features highlights the MAPK signaling pathway and apoptotic pathways.

**Supplementary figure 3:**
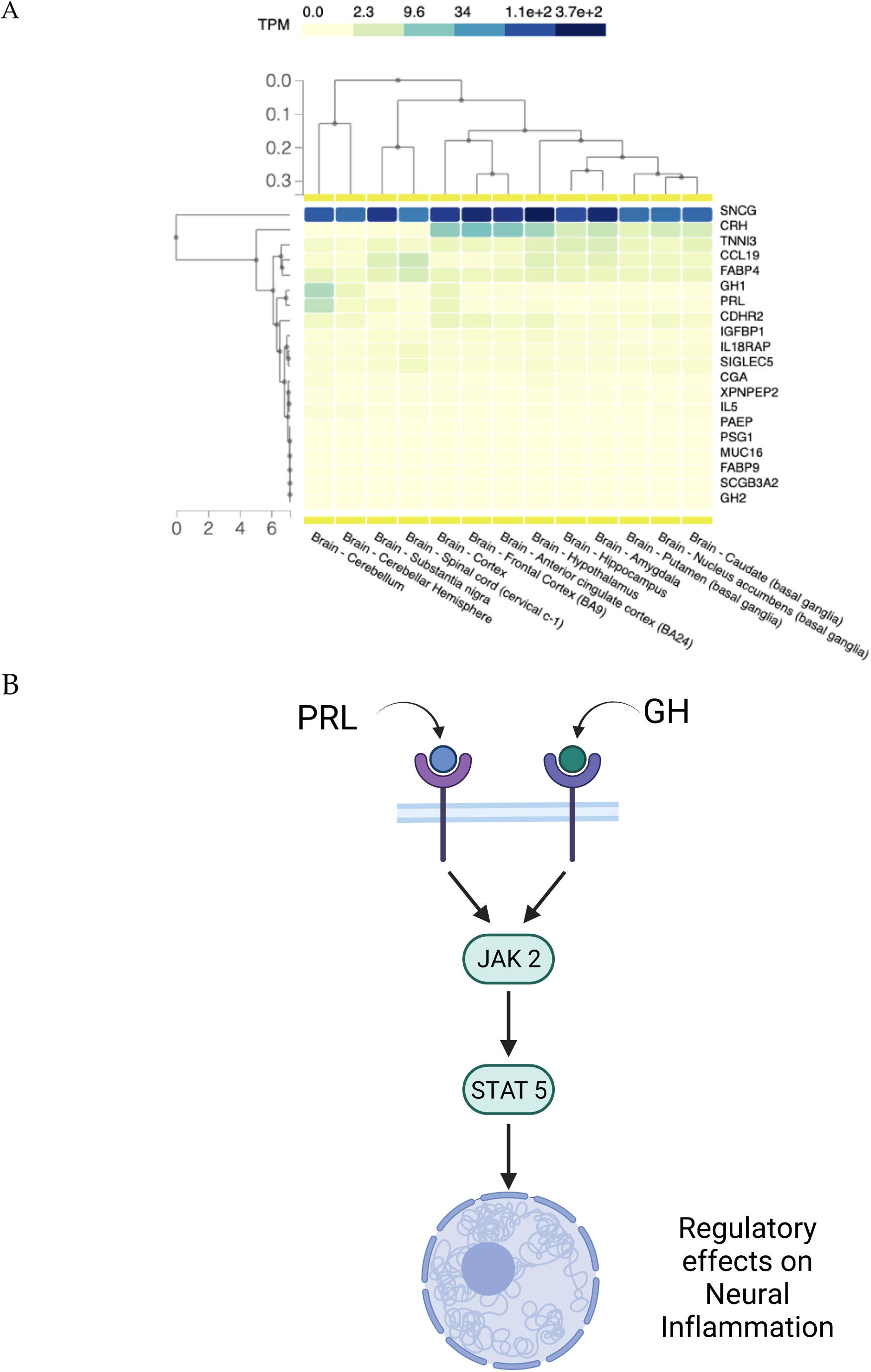
(A) Gene expression levels across different brain tissues from the GTEx study [50], highlighting the differential expression patterns relevant to the pathway including, GH1 and PRL expression in the Cerebellum. (B) The PRL pathway depicting the JAK2-STAT5 signaling cascade, illustrating key molecular interactions and activation steps.

**Supplementary Figure 4:**
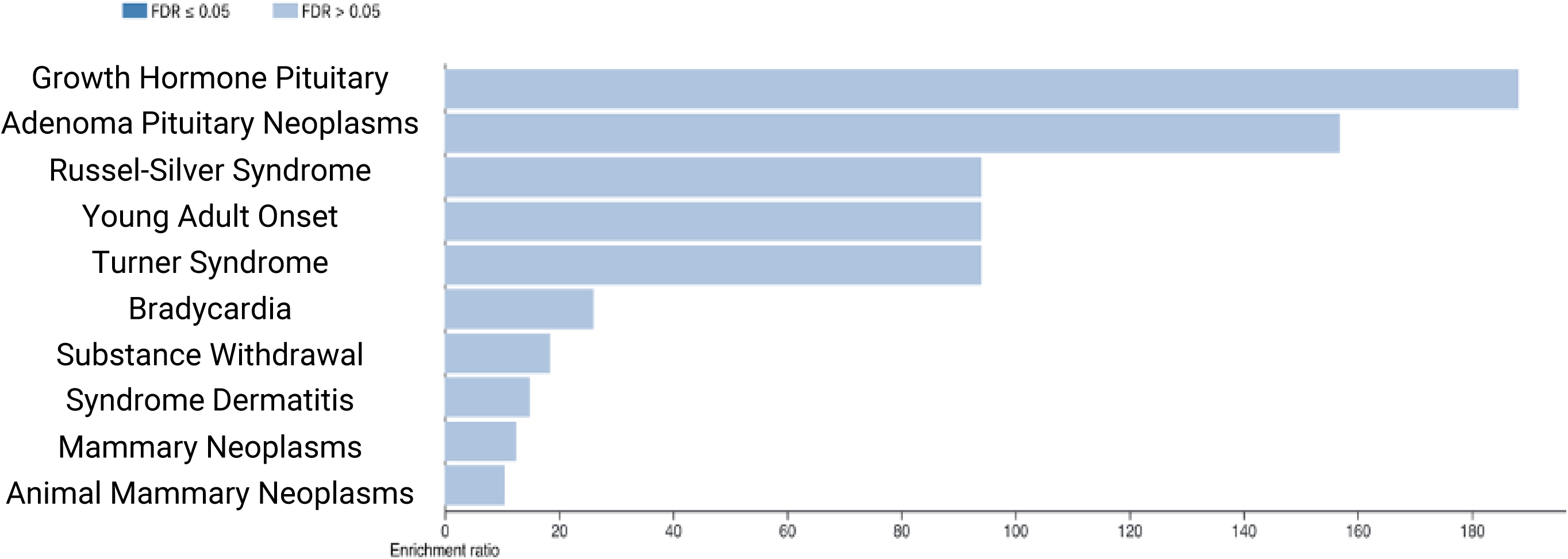
Phenotype-gene enrichment analysis of the top 20 features identified by the neural network model, conducted using the WEB-based GEne SeT AnaLysis Toolkit. The enrichment analysis highlights significant associations between these features and relevant gene sets, providing insights into potential biological pathways linked to the observed phenotypes.

